# Impact of Cardiac Troponin Release and Fluid Resuscitation on Outcomes of Patients with Sepsis

**DOI:** 10.1101/2023.04.04.23288141

**Authors:** Zhiyuan Ma, Mahesh Krishnamurthy, Vivek Modi, David Allen, Jamshid Shirani

## Abstract

**Background:** Septic patients are predisposed to myocardial injury manifested as cardiac troponin release (TnR). Prognostic significance and management implications of TnR and its relationship to fluid resuscitation and outcomes in the intensive care unit (ICU) setting has not been fully elucidated.

**Methods:** A total of 24,778 patients with sepsis from eICU-CRD (n=11,992), MIMIC-III (n=6,638) and MIMIC-IV (n=6,148) databases were included in this retrospective study. In-hospital mortality and survival at 1 year were examined using multivariable logistic, Cox regression analysis, or Kaplan-Meier survival analysis with overlap weighting adjustment, as well as generalized additive models for fluid resuscitation.

**Results:** TnR on admission was associated with higher in-hospital mortality [adjusted odds ratios (OR)=1.33; 95% confidence interval (CI)=1.23-1.43; p<0.001 in unweighted analysis and adjusted OR=1.39; 95% CI=1.29-1.50; p< 0.001 with overlap weighting]. One-year survival was lower in patients with admission TnR (Kaplan-Meier survival analysis; p=0.002). A trend was noted for association between admission TnR and 1-year mortality [adjusted OR=1.16; 95% CI=0.99-1.37; p=0.067 in unweighted analysis] while the association was statistically significant after overlap weighting (adjusted OR=1.25; 95% CI=1.06-1.47; p=0.008). We also found that patients with admission TnR were less likely to benefit from more liberal fluid resuscitation. Adequate fluid resuscitation within a range of 70 to 90 ml/kg in the first 24 hours of ICU stay was associated with lower in-hospital mortality in septic patients without TnR but not in those with admission TnR.

**Conclusions:** Admission TnR was significantly associated with higher in-hospital mortality and worse 1-year survival among septic patients. Adequate fluid resuscitation improved in-hospital mortality in septic patients without but not with admission TnR.

## INTRODUCTION

Patients hospitalized with systemic infection are at risk of developing sepsis-induced myocardial dysfunction or cardiomyopathy, manifested as multiorgan hypoperfusion, cardiac biomarker release, or myocardial dysfunction on imaging [1, 2]. Sepsis-induced myocardial dysfunction has been linked to adverse short and long-term outcomes [3]. Cardiac troponin release (TnR) has correlated well with both systolic and diastolic dysfunction on echocardiography in this context [4]. While short-term adverse impact of TnR has been demonstrated in patients with sepsis [5, 6], the data on long-term impacts of TnR in this setting remains conflicting [7, 8]. The discrepancies may be due to limitations of the prior studies, such as small sample sizes and inadequate adjustment for potential confounding variables.

Aggressive fluid resuscitation to restore tissue perfusion is recommended by the Surviving Sepsis Campaign as the cornerstone of treatment for patient with sepsis [9]. Although more restricted fluid restriction is advocated [10], others have shown evidence against withholding aggressive fluid resuscitation in septic patients with congestive heart failure (CHF) [11, 12].

The aim of this study was to explore the impact of cardiac TnR on short and long-term outcomes of septic patients and its implications for clinical decision making regarding fluid resuscitation strategy by combining data from three large databases and using overlap weighting methods to ensure comparability of between the groups of patients with sepsis and with or without TnR [13].

## METHODS

### Data source

The datasets for investigating the impact of TnR on the outcomes of patients with sepsis were extracted from the Medical Information Mart for Intensive Care III (MIMIC-III) version 1.4, and MIMIC-IV version 1.0 that contain 61,532 and 65,794 ICU stays, respectively.

The databases are publicly available and contain only de-identified health-related data for patients who had been admitted to the critical care units at the Beth Israel Deaconess Medical Center in Boston between 2001 and 2019 [14, 15]. The eICU Collaborative Research Database (eICU-CRD) is also a large database free of protected health information containing 200,859 ICU stays for 139,367 unique patients admitted between 2014 and 2015 to one of the 335 units at 208 hospitals located throughout the United States [16]. The eICU-CRD is completely independent set of data from MIMIC databases, as the source hospital of MIMIC databases did not participate in the eICU program [16]. The database was approved by the Institutional Review Boards of the Massachusetts Institute of Technology. The first author (Z.M.) was given permission to extract data from MIMIC-III, IV and eICU-CRD. The study is conducted in accordance with the Strengthening the Reporting of Observational Studies in Epidemiology reporting guideline. As the databases contain de-identified information, this study was deemed exempt from local institutional research board review.

### Data extraction

Patient variables were extracted, including age, gender, weight, height, ethnicity, comorbidities, in-hospital mortality, Acute Physiology Score (APS) III, and the use of vasopressors and mechanical ventilation in the first 24 hours of the ICU stay. Sequential Organ Failure Assessment (SOFA) score in the first 24 hours of the ICU stay was also extracted as described previously [17]. Cardiac troponin-T was measured in MIMIC-III and IV databases, while troponin-I was measured in eICU-CRD. Admission troponin was defined as the initial measurement of troponin within the first 6 hours of ICU admission. Elevated troponin on admission was defined as a troponin level higher than the upper reference limit value (> 0.04 ng/ml). A significant delta troponin was specified as a rise of troponin by >0.04 ng/ml within 6 hours compared to the admission troponin. Patients who had sepsis and troponin assessment within 6 hours of ICU admission were eligible for inclusion. For patients with multiple ICU admissions, we only included the first ICU stay. We excluded patients who were younger than 18 years old, ICU length of stay less than 1 day or missing values in weight, gender, or SOFA scores.

### Definition of sepsis

Sepsis was defined according to the Third International Consensus Definitions for Sepsis and Septic Shock (Sepsis-3) [18]. Suspected infection was identified if the collection of a body fluid culture temporally prior to administration of antibiotics occurred from MIMC-III and IV [17]. We could not extract suspected infection from eICU database. Instead, a bacterial or fungal infection was sourced from ICD-9 codes for infection as described in the Appendix 1 of the cited paper [3].

### Statistical analysis

The Primary outcome of the study was in-hospital mortality. The secondary outcome was 1-year survival. To investigate the potential impact of troponin elevation on the primary outcome, multivariable regression analysis was applied. Clinically relevant confounders including age, body mass index (BMI), gender, ethnicity, SOFA score, and interventions (mechanical ventilation and vasopressor use) were entered into a multivariate logistic regression model as covariates. The variable “hospital” was also included in the analyses for eICU-CRD and integrating data for overall effects to reduce biases resulting from differences among hospitals. BMI was derived from height and weight. Missing values for height was imputed with a multivariable linear regression model, specifically, weight, ethnicity, and gender were used for the imputation of the height variable. Multivariable Cox regression analysis was performed to derive hazard ratios, which were stratified by age and SOFA score and adjusted for BMI and gender. To account for the potential confounders associated with the troponin elevation and to ensure the robustness of our results, overlap weighting was used based on propensity scores [13]. Propensity scores for the probability of the elevation of troponin on admission were estimated from a multivariable logistic regression model containing age, BMI, gender, ethnicity, comorbidities [CHF, coronary artery disease (CAD), asthma, chronic obstructive pulmonary disease (COPD), atrial fibrillation (AF), chronic renal disease, chronic liver disease, and stroke]. Sensitivity analyses were performed using APSIII instead of SOFA as a covariable. Subgroup analyses were performed in patients with delta troponin values and patients with fluid resuscitation in the first 24 hours of ICU stay. Survival analyses were performed in MIMIC-III that has extractable survival data. Fluid resuscitation analysis was only performed in MIMIC-III and IV databases, as we could not extract the intravenous fluid volume from eICU-CRD due to lack of database structure. Given that restrictive fluid resuscitation and liberal fluid resuscitation in sepsis are associated with worse outcomes, there is likely a nonlinear relationship between fluid resuscitation and in-hospital mortality. To account for such nonlinearity, generalized additive models were used to assess the association between fluid resuscitation and in-hospital mortality adjusting for age, gender, ethnicity, BMI, SOFA on the first day of ICU stay, use of vasopressors and use of mechanical ventilation. All continuous variables were included as having potentially nonlinear associations.

Data were presented as mean and 95% confidence interval (CI) for continuous variables. For differences in continuous variables, Student’s t-test (normally distributed) was used for comparison. Categorical variables were expressed as percentages (%) and were compared using χ^2^ test. The risk of primary outcome was calculated as odds ratio (OR) using multivariable logistic regression models. Log-rank test was used to test survival differences. P values <0.05 were considered statistically significant. All analyses were conducted with R statistical analysis software version 4.1.2. (Vienna, Austria. https://www.R-project.org/).

## RESULTS

### Basic characteristics of study population

A total of 11,992, 6,638 and 6,148 septic patients with admission troponin were enrolled in the final cohorts based on study criteria (Figure 1) from the 200,859, 61,532 and 65,794 ICU stays in eICU-CRD, MIMIC-III and MIMIC-IV, respectively. Number of patients with normal and elevated troponin levels were 5,735 and 6, 257 in eICU-CRD, 1,766 and 4,872 in MIMC-III, and 1,732 and 4,416 in MIMIC-IV. In-hospital mortality rates were 14.9%, 17.4%, 22.1% in the normal troponin groups, and 24.2%, 22.3% and 29.4% for the elevated troponin groups in eICU-CRD, MIMIC-III and MIMIC-IV, respectively. Mean SOFA score was higher in septic patients with TnR on admission compared to those with normal troponin (Table 1). The baseline characteristics of these cohorts were summarized in Table 1 and Supplementary table 1-3.

**Figure 1.**
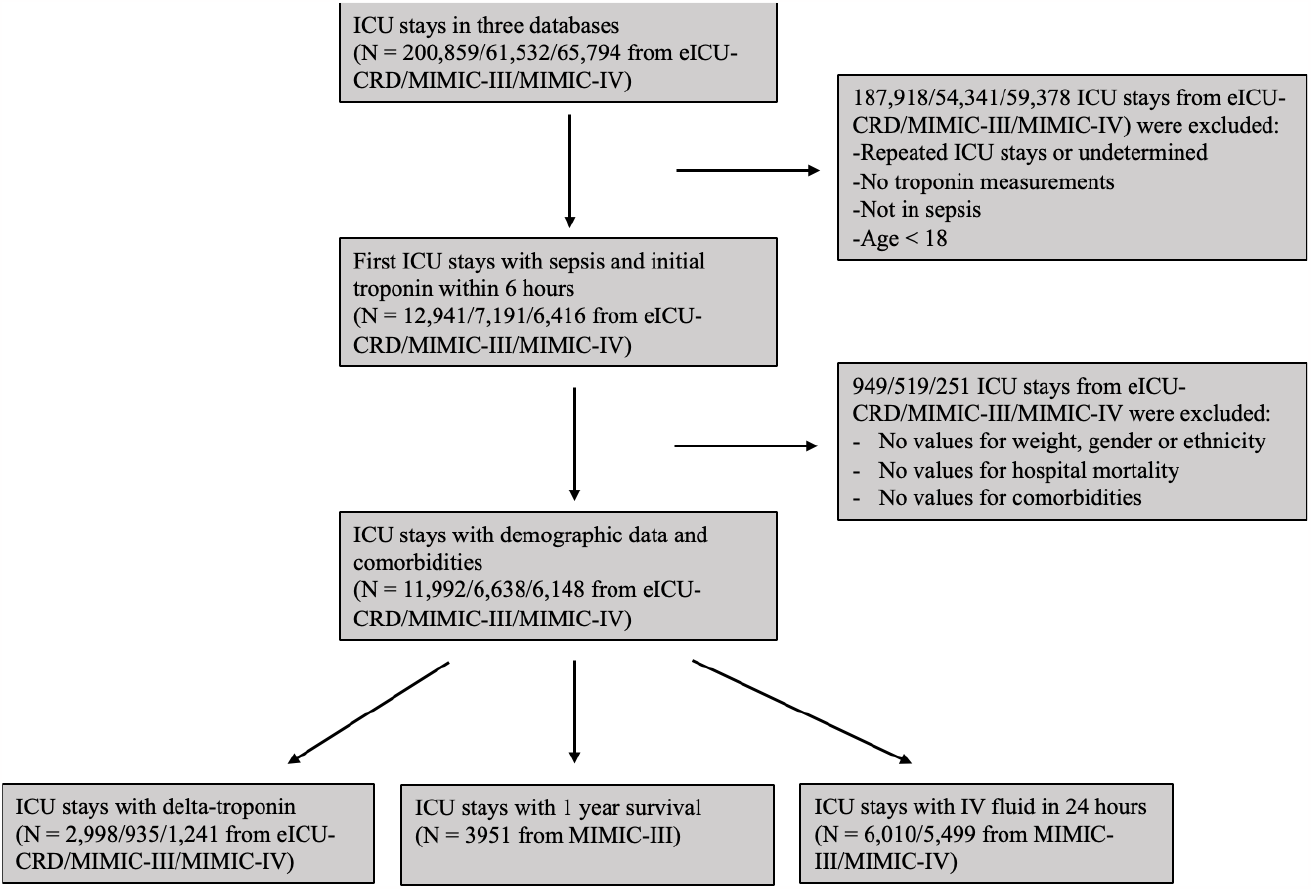
Flowchart illustrating the process of selection of intensive care stays for sepsis.

**Table 1.** Comparisons of baseline characteristics of between patients with normal troponin and those with elevated troponin on admission.

| Variables | Unweighted |  |  | OW |  |  |
| --- | --- | --- | --- | --- | --- | --- |
|  | Normal troponin | Elevated troponin | P value* | Normal troponin | Elevated troponin | P value* |
| **eICU-CRD** |  |  |  |  |  |  |
| N | 5735 | 6257 |  | 2878.9 | 2878.9 |  |
| Age (Mean ± SD) | 66.6 (15.2) | 70.5 (14.5) | <0.001 | 68.6 (14.6) | 68.6 (15.0) | 1.000 |
| Male – no. (%) | 3004 (52.4) | 3338 (53.3) | 0.297 | 1521.1 (52.8) | 1521.1 (52.8) | 1.000 |
| BMI (Mean ± SD) | 29.7 (9.6) | 28.8 (8.6) | <0.001 | 29.3 (9.1) | 29.3 (9.0) | 1.000 |
| Mech – no. (%) | 2460 (42.9) | 2988 (47.8) | <0.001 | 1219.4 (42.4) | 1396.5 (48.5) | <0.001 |
| Vaso – no. (%) | 1199 (20.9) | 1755 (28.0) | <0.001 | 604.4 (21.0) | 814.0 (28.3) | <0.001 |
| SOFA (Mean ± SD) | 6.8 (3.5) | 7.8 (3.7) | <0.001 | 6.8 (3.5) | 7.8 (3.8) | <0.001 |
| APSIII (Mean ± SD) | 66.5 (26.3) | 74.8 (28.8) | <0.001 | 67.4 (26.1) | 74.3 (29.1) | <0.001 |
| Mortality – no. (%) | 854 (14.9) | 1512 (24.2) | <0.001 | 445.3 (15.5) | 674.9 (23.4) | <0.001 |
| **MIMIC-III** |  |  |  |  |  |  |
| N | 1766 | 4872 |  | 1232.0 | 1232.0 |  |
| Age (Mean ± SD) | 73.5 (14.2) | 71.0 (15.1) | <0.001 | 72.9 (14.3) | 72.9 (14.5) | 1.000 |
| Male – no. (%) | 953 (54.0) | 2795 (57.4) | 0.015 | 670.3 (54.4) | 670.3 (54.4) | 1.000 |
| BMI (Mean ± SD) | 28.1 (7.6) | 28.0 (7.1) | 0.654 | 28.0 (7.5) | 28.0 (7.4) | 1.000 |
| Mech – no. (%) | 779 (44.1) | 2387 (49.0) | <0.001 | 542.3 (44.0) | 630.9 (51.2) | <0.001 |
| Vaso – no. (%) | 545 (30.9) | 1826 (37.5) | <0.001 | 380.0 (30.8) | 468.5 (38.0) | <0.001 |
| SOFA (Mean ± SD) | 5.0 (3.3) | 5.5 (3.5) | <0.001 | 5.0 (3.3) | 5.7 (3.5) | <0.001 |
| APSIII (Mean ± SD) | 50.5 (20.9) | 52.7 (22.2) | <0.001 | 50.2 (20.8) | 54.4 (22.4) | <0.001 |
| Mortality – no. (%) | 308 (17.4) | 1085 (22.3) | <0.001 | 207.9 (16.9) | 308.8 (25.1) | <0.001 |
| **MIMIC-IV** |  |  |  |  |  |  |
| N | 1732 | 4416 |  | 1194.1 | 1194.1 |  |
| Age (Mean ± SD) | 70.7 (15.0) | 69.1 (15.3) | <0.001 | 70.3 (15.1) | 70.3 (15.0) | 1.000 |
| Male – no. (%) | 916 (52.9) | 2635 (59.7) | <0.001 | 652.7 (54.7) | 652.7 (54.7) | 1.000 |
| BMI (Mean ± SD) | 28.5 (7.8) | 28.3 (7.1) | 0.467 | 28.4 (7.7) | 28.4 (7.4) | 1.000 |
| Mech – no. (%) | 736 (42.5) | 1948 (44.1) | 0.262 | 504.7 (42.3) | 533.3 (44.7) | 0.095 |
| Vaso – no. (%) | 850 (49.1) | 2692 (61.0) | <0.001 | 585.8 (49.1) | 728.8 (61.0) | <0.001 |
| SOFA (Mean ± SD) | 3.6 (2.0) | 4.0 (2.2) | <0.001 | 3.6 (2.0) | 4.0 (2.2) | <0.001 |
| APSIII (Mean ± SD) | 62.5 (25.7) | 66.5 (27.0) | <0.001 | 62.1 (25.5) | 67.3 (27.2) | <0.001 |
| Mortality – no. (%) | 382 (22.1) | 1300 (29.4) | <0.001 | 261.4 (21.9) | 370.5 (31.0) | <0.001 |

APSIII=acute physiology score III; Mech=mechanical ventilation; OW=overlap weighting; SOFA= sequential organ failure assessment score; Vaso=vasopressor use.

### Admission troponin and in-hospital mortality

Multivariable logistic regression analyses, adjusted for age, BMI, gender, SOFA score, mechanical ventilation, and vasopressor use, showed that compared to normal troponin, TnR on admission was significantly associated with higher in-hospital mortality in eICU-CRD (adjusted OR=1.33; 95% CI=1.18-1.46; p< 0.001), MIMIC-III (adjusted OR=1.25; 95% CI=1.07-1.46; p=0.004) and MIMIC-IV (adjusted OR=1.30; 95% CI=1.14-1.50; p<0.001) (Figure 2). To account for potential confounding variables, the impact of TnR on outcomes was also evaluated using multivariable logistic regression analyses after propensity score-based overlap weighting. The findings were similar with odd ratios of 1.32 (95% CI=1.19-1.47; p<0.001) in eICU-CRD, 1.37 (95% CI=1.17-1.60; p<0.001) in MIMIC-III, and 1.39 (95% CI=1.20-1.60; p<0.001) in MIMIC-IV. The adverse impact of TnR on in-hospital mortality was also reproduced when the 3 databases were integrated with an overall adjusted OR of 1.33 (95% CI=1.23-1.43; p<0.001) in the unweighted analysis and 1.39 (95% CI=1.29-1.50; P<0.001) with overlap weighting (Figure 2).

**Figure 2.**
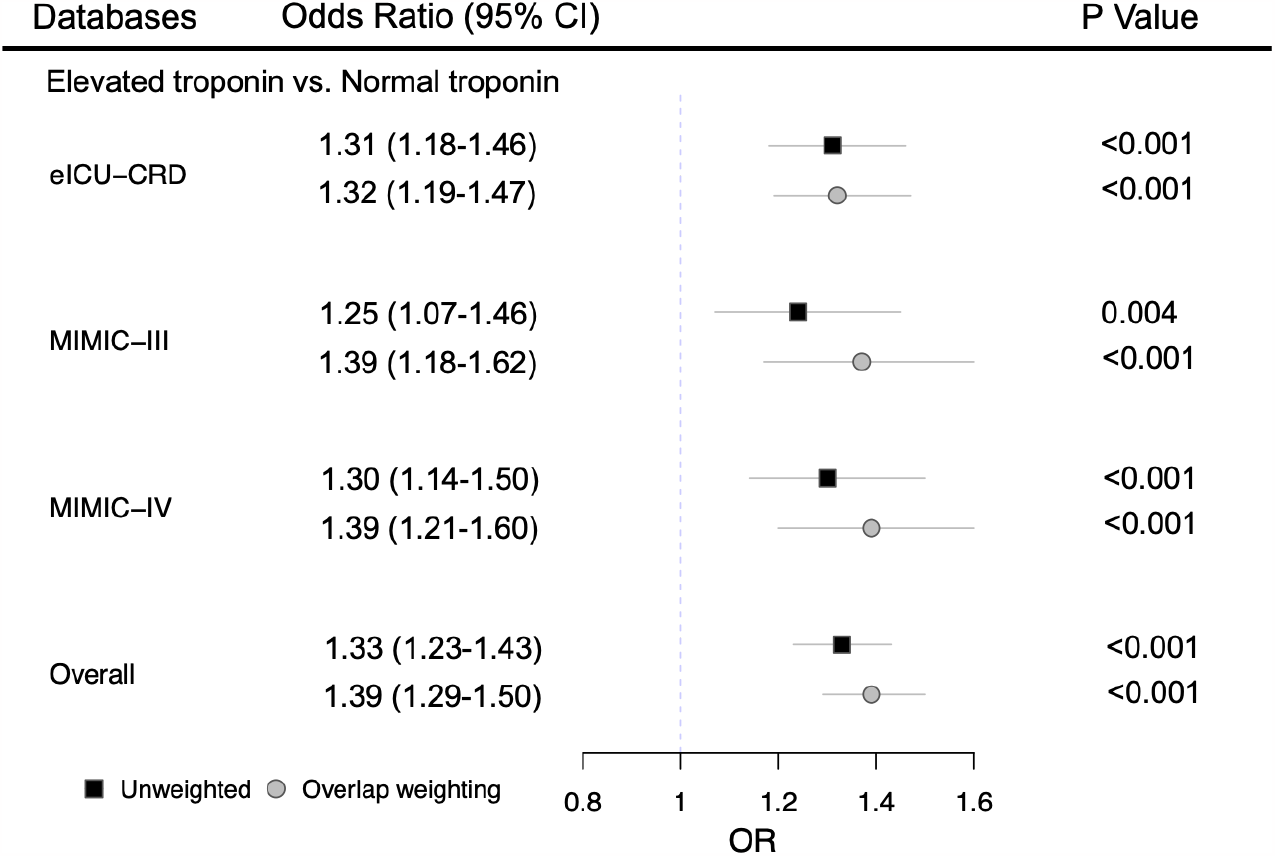
Forest plot demonstrating the uniform and direct association of admission cardiac troponin release and in-hospital mortality in three datasets.

### TnR and 1-year survival

To address the association between TnR and long-term survival, we performed Kaplan-Meier survival analyses for patients in the MIMIC-III database. As shown in Figure 3, both unadjusted and adjusted 1-year survivals were lower in patients with TnR (p=0.002). In the multivariable logistic regression analyses, there was a trend towards higher 1-year mortality in those with TnR (adjusted OR=1.16; 95% CI=0.99-1.37; p=0.067), that became statistically significant after overlap weighting (adjusted OR=1.25; 95% CI=1.06-1.47; p=0.008). The reliability of the results was further tested by using Cox proportional hazards models after adjusting for confounding variables. While adjusted hazard ratio for the association between TnR and 1-year mortality was 1.19 (95% CI=0.95-1.48; p=0.123), in the unweighted analysis, the association was again statistically significant [hazard ratio=1.24 (95% CI=1.01-1.54; p=0.041) after overlap weighting.

**Figure 3.**
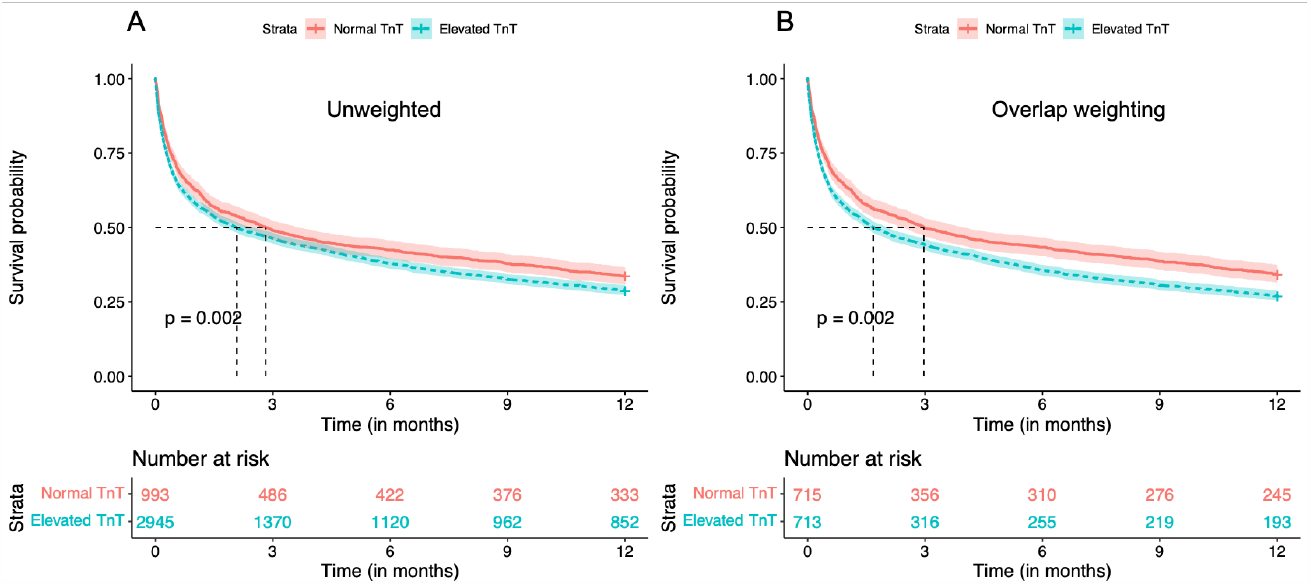
One-year survival in septic patients with or without troponin elevation in MIMC-III. (A) 1-year survival analysis without propensity scored-based overlap weighting. (B) 1-year survival analysis with propensity scored-based overlap weighting. TnT=troponin-T.

### Delta troponin and in-hospital mortality

To determine whether serial troponin measurements play a role in the management of sepsis, we assessed the relationship between delta troponin and in-hospital mortality. In the unweighted multivariable logistic regression analyses, there was a trend towards an association between increased in-hospital mortality and delta troponin in eICU-CRD (adjusted OR=1.22; 95% CI=1.00-1.58; p=0.054) while no such association was observed in MIMIC-III (adjusted OR=1.12; 95% CI=0.77-1.63; p=0.541). On the contrary, delta troponin was found to be significantly associated with higher in-hospital mortality [adjust OR=1.38 (95% CI=1.06-1.79; p=0.017)] in MIMIC-IV (Figure 4). Similar results were achieved in the analyses with overlap weighting. Furthermore, we found that delta troponin was significantly associated with higher in-hospital mortality when pooled data from the three databases was used for both unweighted [adjusted OR=1.29 (95% CI=1.10-1.51; p=0.001)] and weighted [adjusted OR=1.36 (95% CI=1.16-1.59; p<0.001)] (Figure 4).

**Figure 4.**
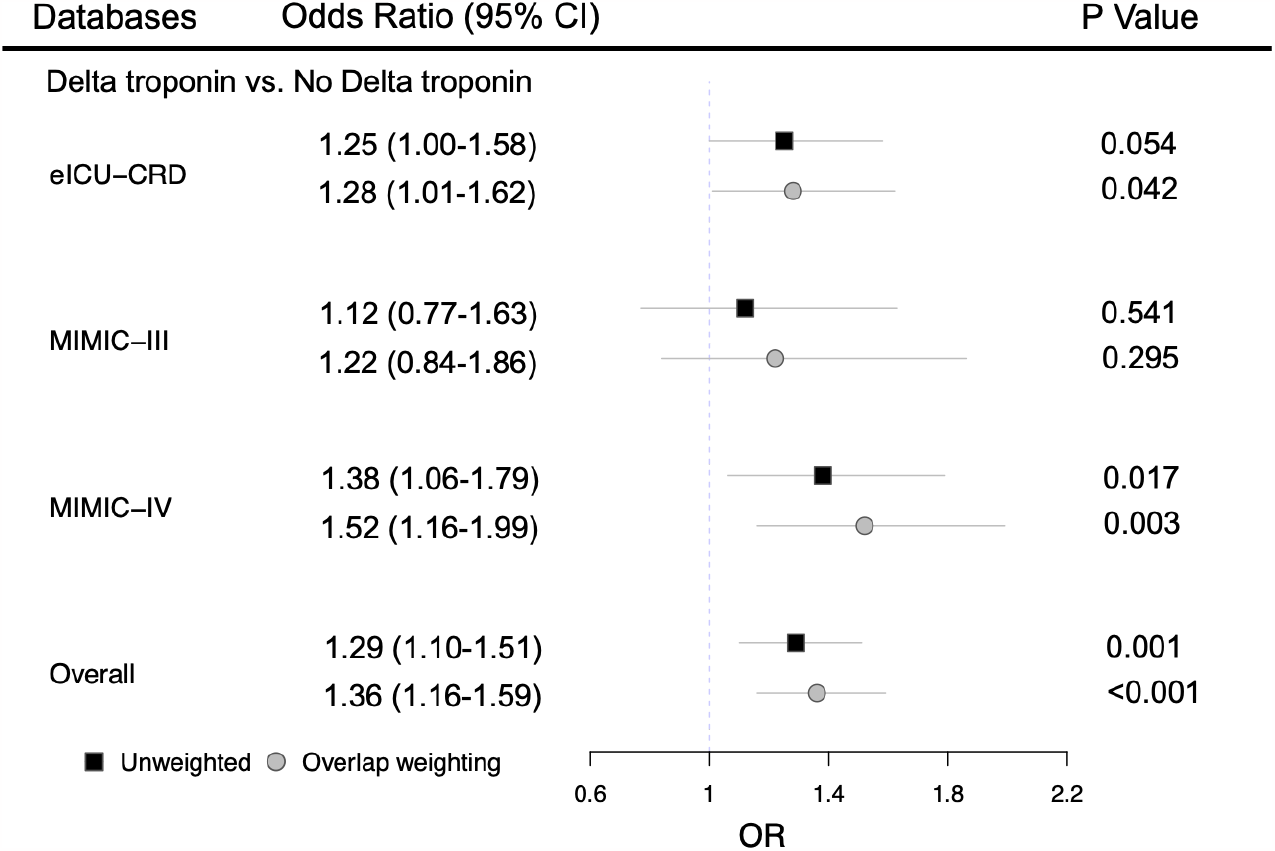
Forest plot demonstrating the impact of admission delta cardiac troponin release and in-hospital mortality separately in the three datasets and in the overall (pooled) data for unweighted and weighted analyses.

### Fluid resuscitation in septic patients with TnR

Fluid resuscitation in patients with sepsis is considered the cornerstone of treatment [9]. We performed nonlinear regression using generalized additive models to analyze the association between intravenous fluid resuscitation, TnR and in-hospital mortality. The data indicated that there was a nearly U-shaped association between fluid resuscitation and in-hospital mortality in patients with or without TnR in both MIMIC-III and MIMIC-IV databases (Figure 5). The highest in-hospital mortality occurred in patients with fluid resuscitation of 30-40 ml/kg during the first 24 hours of ICU stay, whereas restrictive and liberal fluid resuscitation resulted in reduced mortality. Fluid resuscitation of >40 ml/kg was associated with higher in-hospital mortality in those with TnR while it resulted in improved survival in those without elevated troponin with the highest survival noted among patients receiving fluid resuscitation at a range of 70 to 90 ml/kg (Figure 5).

**Figure 5.**
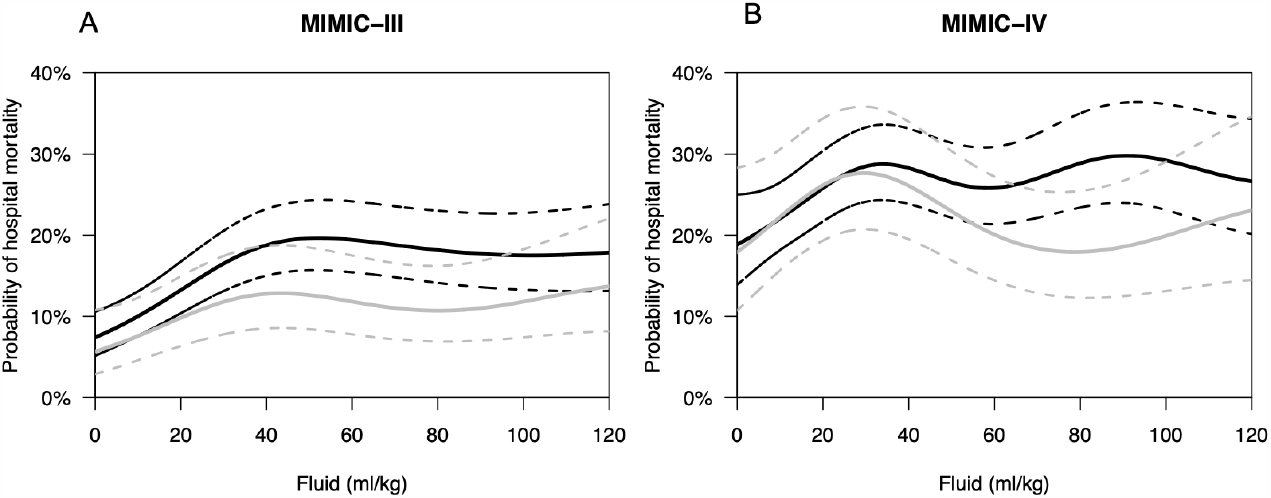
Probability of hospital mortality with intravenous fluid resuscitation. The association of in-hospital mortality and fluid resuscitation during the first 24 hours of intensive care unit (ICU) stay is visually summarized from the generalized additive model fit on MIMIC-III (A) and from MIMIC-IV (B) data after adjustment for age, gender, ethnicity, body mass index, SOFA score on the first day of the ICU stay, the use of vasopressors and mechanical ventilation. The solid line is the mean prediction, and the dashed lines are the 95% confidence intervals. Back line denotes septic patients with elevated troponin on admission. Grey line represents septic patients without troponin elevation.

### Sensitivity analyses

To verify the robustness of the models, we performed sensitivity analyses using APSIII scores instead of SOFA scores as covariables. The sensitivity analyses for TnR and delta troponin (Supplementary table 4), 1-year mortality (Supplementary table 5), and intravenous fluid resuscitation (Supplementary figure 1) were consistent with those obtained with SOFA scores.

## Discussion

Myocardial dysfunction has been reported frequently in the setting of sepsis and septic shock [1, 19]. The precise mechanisms of cardiac dysfunction and myocyte injury are incompletely understood and likely multi-faceted and multi-factorial. One set of processes leading to myocyte injury is through a cascade of events initiated with the systemic inflammatory response to infection leading to immune activation, release of pro-inflammatory cytokines, oxidative stress, endothelial damage and dysfunction leading to myocyte injury and cell death [2, 20]. Clinically, troponin-I and troponin-T are well established markers for cardiac myocyte injury and TnR in the setting of sepsis has been shown to be associated with adverse outcomes [5, 6]. In a retrospective study of 944 patients with severe sepsis and septic shock, logarithm-transformed troponin-T was directly associated with in-hospital mortality (OR=1.4, p=0.04) [7]. In another prospective study of 1124 patients with sepsis, adjusted Cox regression analysis showed a direct association between elevated high-sensitivity troponin I and increased mortality within the first 14 days [8]. In line with these findings, we found that TnR was significantly associated with worse in-hospital mortality in patients with sepsis using real-world large databases. These results provide convincing evidence that TnR is associated with high short-term mortality in patient with sepsis.

Serial testing to trend troponin is a common clinical practice. However, the role of such serial testing has not been well studied in the setting of sepsis. In this study, we assessed the clinical importance of serial troponin testing by probing the relationship between delta troponin in patients with sepsis and in-hospital mortality. We found discrepant results in association of delta troponin and in-hospital mortality in the individual databases used in this study with no association in MIMIC-III, a trend toward an association in eICU-CRD, and a significant association in MIMIC-IV. However, a significant association was found between delta troponin and in-hospital mortality when the pooled data was analyzed. Others have reported an association between delta troponin T and in-hospital mortality only in unadjusted multivariable analysis [7]. There are significant methodological differences in the type and timing of troponin measurements among studies and thus, further studies are needed to investigate the role of serial troponin testing in sepsis.

There is conflicting evidence on the association of TnR with long-term mortality and complications. It has been reported that admission troponin-T was associated with 1-year mortality in unadjusted analysis [7]. We found that both unadjusted and adjusted 1-year survival was lower in patients with TnR in the MIMIC-III database. In patients with end-stage renal disease, TnR was associated with increased mortality up to 4 years after the index admission for sepsis [21]. Moreover, a retrospective analysis of patients without preexisting cardiovascular disease revealed that TnR during sepsis was associated with an increased risk of cardiovascular complications beyond 1 year [22]. An association between TnR (high-sensitivity troponin I) and long-term (beyond 14 days) mortality was not shown in an adjusted Cox regression analysis in a prospective study [8]. In this study, we found that there was a trend for an association between TnR and 1-year mortality in multivariable logistic and Cox regression analyses, and a significantly associated between them with overlap weighting in MIMIC-III. Future studies are needed to determine the association between TnR and long-term mortality in patients with sepsis.

The role of early and continued fluid resuscitation in sepsis and septic shock remains controversial. While early fluid resuscitation has been adopted in general as a means of improving hemodynamics and stroke volume, universal intravenous fluid administered at a rate of 98 to 130 ml/kg within the first 72 hours of admission has failed to improve outcomes in several clinical trials [23-25]. It is yet to be determined whether subsets of patients can be identified who would benefit from, or otherwise be harmed by, routine early fluid resuscitation. In this study we have identified a subset of patients (with TnR) who in fact have higher in-hospital mortality when receiving large amounts of fluid intravenously. In-hospital mortality was thus higher in patients with TnR who received fluids at a rate of >90 ml/kg in the first 24 hours of intensive care unit stay. Others have also reported that in-hospital mortality could be the lowest if fluid resuscitation in the setting of sepsis does not exceed ∼5 liters within the first 24 hours [26]. We can only speculate about the pathophysiology of the negative impact of fluid resuscitation in sepsis associated with TnR. Interstitial and cellular edema are frequently observed in sepsis and septic shock. The heart is especially susceptible to myocardial edema due to its dense capillary network and high interstitial flow rate [27]. Recently, myocardial edema has been recognized as a major contributor to cardiac dysfunction in patients with sepsis [28]. It is thus plausible that, in the presence of early cardiac involvement in sepsis, aggressive early fluid resuscitation may exacerbate myocardial edema and potentially lead to myocardial ischemia due to microvascular obstruction.

### Study limitations

Several limitations are present in our study. First, Missing values for height was imputed using multivariable linear regression. Errors could have been introduced in the study despite this generally reliable imputation strategy. Second, although adjustments for available potential confounding variables were made, other variables may have existed but not available to adjustment due to the retrospective nature of the study. Finally, the results reported for the long-term mortality and fluid resuscitation may not be generalized due to data being limited to one of the three databases used.

## Conclusion

In conclusion, our finding demonstrated that TnR was significantly associated with higher in-hospital mortality and that 1-year survival for septic patients with TnR was lower than those without troponin release. We also found that aggressive fluid resuscitation improved in-hospital mortality in septic patients without but not with TnR.

## Data Availability

eICU-CRD is available from https://eicu-crd.mit.edu. MIMIC-III and IV are available from https://mimic.mit.edu. eICU-CRD, MIMIC-III and IV datasets are not owned or collected by any of the authors.

## Author Contributions

ZM and JS designed the study and wrote the manuscript. ZM performed and analyzed data. JS supervised the research. MK, VM, DA and JS reviewed data and revised the manuscript. All authors reviewed the results and approved the final version of the manuscript.

