## Supplemental figure and tables for "Impact of Cardiac Troponin Release and Fluid Resuscitation on Outcomes of Patients with Sepsis"

Short running title: Ma: Cardiac TnR in Sepsis

Zhiyuan Ma MD^1^, Mahesh Krishnamurthy MD^1^, Vivek Modi MD^2^, David Allen DO^2^, Jamshid Shirani, MD^2^

From the Departments of Internal Medicine^1^ and Cardiology^2^, St. Luke’s University Health Network, Bethlehem, PA

Address for correspondence: Zhiyuan Ma or Jamshid Shirani, St. Luke’s University Health Network, 801 Ostrum Street, Bethlehem, PA 18015. Telephone: 484-526-4011; FAX: 484-526-4010; (ZM); (JS)

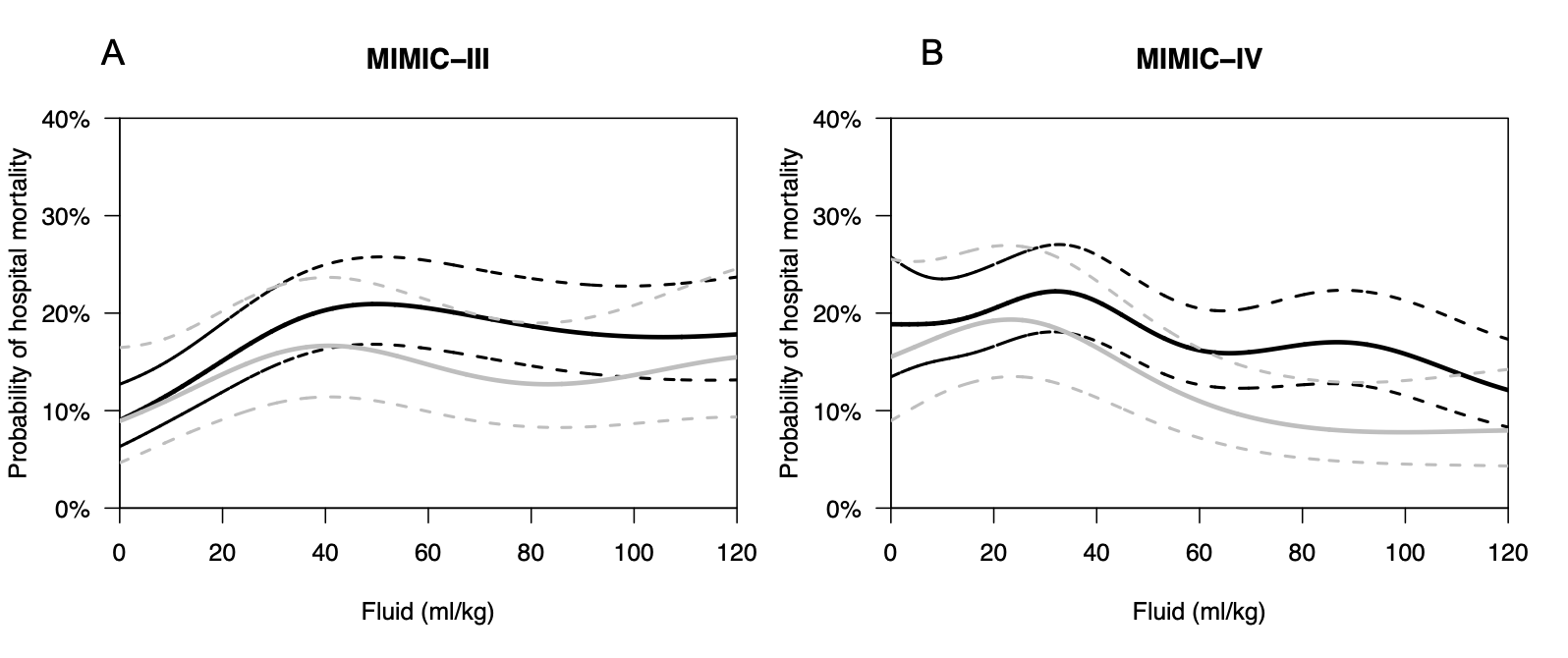

Supplementary figure 1. Probability of hospital mortality with intravenous fluid resuscitation. The association of in-hospital mortality and fluid resuscitation during the first 24 hours of intensive care unit (ICU) stay is visually summarized from the generalized additive model fit on MIMIC-III (A) and from MIMIC-IV (B) after the adjustment for age, gender, ethnicity, body mass index, acute physiology score III on the first day of the ICU stay, the use of vasopressors and mechanical ventilation. The solid line is the mean prediction, and the dashed lines are the 95% CIs. Back line denotes septic patients with elevated troponin on admission. Grey line represents septic patients without troponin elevation.

Supplementary Table 1. Demographic and clinical characteristics before and after propensity score-based overlap weighting in eICU-CRD data.

|  | Unweighted | | | OW | | |
| --- | --- | --- | --- | --- | --- | --- |
| Variables | Normal troponin | Elevated troponin | SMD | Normal troponin | Elevated troponin | SMD |
| N | 5735 | 6257 |  | 2878.9 | 2878.9 |  |
| Age (Mean ± SD) | 66.6 (15.2) | 70.5 (14.5) | 0.263 | 68.6 (14.6) | 68.6 (15.0) | <0.001 |
| Male – no. (%) | 3004 (52.4) | 3338 (53.3) | 0.019 | 1521.1 (52.8) | 1521.1 (52.8) | <0.001 |
| Ethnicity (%) |  |  | 0.086 |  |  | <0.001 |
| African American | 447 (7.8) | 568 (9.1) |  | 238.1 (8.3) | 238.1 (8.3) |  |
| Asian | 79 (1.4) | 128 (2.0) |  | 48.2 (1.7) | 48.2 (1.7) |  |
| Hispanic | 285 (5.0) | 270 (4.3) |  | 132.6 (4.6) | 132.6 (4.6) |  |
| Native American | 42 (0.7) | 29 (0.5) |  | 17.4 (0.6) | 17.4 (0.6) |  |
| Other/Unknown | 297 (5.2) | 288 (4.6) |  | 142.7 (5.0) | 142.7 (5.0) |  |
| White | 4585 (79.9) | 4974 (79.5) |  | 2299.9 (79.9) | 2299.9 (79.9) |  |
| BMI (Mean ± SD) | 29.7 (9.6) | 28.8 (8.6) | 0.104 | 29.3 (9.1) | 29.3 (9.0) | <0.001 |
| CHF – no. (%) | 741 (12.9) | 1329 (21.2) | 0.222 | 466.5 (16.2) | 466.5 (16.2) | <0.001 |
| CKD – no. (%) | 648 (11.3) | 1123 (17.9) | 0.189 | 401.0 (13.9) | 401.0 (13.9) | <0.001 |
| CLD – no. (%) | 101 (1.8) | 100 (1.6) | 0.013 | 47.9 (1.7) | 47.9 (1.7) | <0.001 |
| COPD – no. (%) | 870 (15.2) | 832 (13.3) | 0.054 | 407.5 (14.2) | 407.5 (14.2) | <0.001 |
| CAD – no. (%) | 70 (1.2) | 136 (2.2) | 0.074 | 44.9 (1.6) | 44.9 (1.6) | <0.001 |
| Stroke – no. (%) | 112 (2.0) | 152 (2.4) | 0.033 | 63.7 (2.2) | 63.7 (2.2) | <0.001 |

BMI=body mass index; CHF=congestive heart failure; CAD=coronary artery disease; CKD=chronic kidney disease; CLD=chronic liver disease; COPD=chronic obstructive pulmonary disease; OW= overlap weighting; SMD, standardized mean difference.

Supplementary Table 2. Demographic and clinical characteristics before and after propensity score-based overlap weighting in MIMIC-III.

|  | Unweighted | | | OW | | |
| --- | --- | --- | --- | --- | --- | --- |
| Variables | Normal troponin | Elevated troponin | SMD | Normal troponin | Elevated troponin | SMD |
| N | 1766 | 4872 |  | 1232.0 | 1232.0 |  |
| Age (Mean ± SD) | 73.5 (14.2) | 71.0 (15.1) | 0.166 | 72.9 (14.3) | 72.9 (14.5) | <0.001 |
| Male – no. (%) | 953 (54.0) | 2795 (57.4) | 0.069 | 670.3 (54.4) | 670.3 (54.4) | <0.001 |
| Ethnicity (%) |  |  | 0.205 |  |  | <0.001 |
| African American | 160 (9.1) | 374 (7.7) |  | 108.7 (8.8) | 108.7 (8.8) |  |
| Asian | 54 (3.1) | 102 (2.1) |  | 33.8 (2.7) | 33.8 (2.7) |  |
| Hispanic | 37 (2.1) | 108 (2.2) |  | 27.1 (2.2) | 27.1 (2.2) |  |
| Native American | 1 (0.1) | 3 (0.1) |  | 0.7 (0.1) | 0.7 (0.1) |  |
| Other/Unknown | 183 (10.4) | 832 (17.1) |  | 143.8 (11.7) | 143.8 (11.7) |  |
| White | 1331 (75.4) | 3453 (70.9) |  | 917.8 (74.5) | 917.8 (74.5) |  |
| BMI (Mean ± SD) | 28.1 (7.6) | 28.0 (7.1) | 0.012 | 28.0 (7.5) | 28.0 (7.4) | <0.001 |
| Afib – no. (%) | 680 (38.5) | 1652 (33.9) | 0.096 | 459.4 (37.3) | 459.4 (37.3) | <0.001 |
| CHF – no. (%) | 460 (26.0) | 1367 (28.1) | 0.045 | 327.5 (26.6) | 327.5 (26.6) | <0.001 |
| CKD – no. (%) | 341 (19.3) | 1051 (21.6) | 0.056 | 249.3 (20.2) | 249.3 (20.2) | <0.001 |
| CLD – no. (%) | 99 (5.6) | 157 (3.2) | 0.116 | 60.2 (4.9) | 60.2 (4.9) | <0.001 |
| COPD – no. (%) | 62 (3.5) | 144 (3.0) | 0.031 | 41.4 (3.4) | 41.4 (3.4) | <0.001 |
| CAD – no. (%) | 439 (24.9) | 2153 (44.2) | 0.415 | 358.5 (29.1) | 358.5 (29.1) | <0.001 |
| Stroke – no. (%) | 213 (12.1) | 464 (9.5) | 0.082 | 140.4 (11.4) | 140.4 (11.4) | <0.001 |

AFIB=atrial fibrillation; all other abbreviations as in Supplementary Table 1.

Supplementary table 3. Demographic and clinical characteristics before and after propensity score-based overlap weighting in MIMIC-IV.

|  | Unweighted | | | OW | | |
| --- | --- | --- | --- | --- | --- | --- |
| Variables | Normal troponin | Elevated troponin | SMD | Normal troponin | Elevated troponin | SMD |
| N | 1732 | 4416 |  | 1194.11 | 1194.11 |  |
| Age (Mean ± SD) | 70.7 (15.0) | 69.1 (15.3) | 0.107 | 70.3 (15.1) | 70.3 (15.0) | <0.001 |
| Male – no. (%) | 916 (52.9) | 2635 (59.7) | 0.137 | 652.7 (54.7) | 652.7 (54.7) | <0.001 |
| Ethnicity (%) |  |  | 0.107 |  |  | <0.001 |
| African American | 191 (11.0) | 446 (10.1) |  | 128.6 (10.8) | 128.6 (10.8) |  |
| Asian | 55 (3.2) | 108 (2.4) |  | 34.8 (2.9) | 34.8 (2.9) |  |
| Hispanic | 51 (2.9) | 119 (2.7) |  | 34.8 (2.9) | 34.8 (2.9) |  |
| Native American | 2 (0.1) | 9 (0.2) |  | 1.6 (0.1) | 1.6 (0.1) |  |
| Other/Unknown | 305 (17.6) | 946 (21.4) |  | 223.1 (18.7) | 223.1 (18.7) |  |
| White | 1128 (65.1) | 2788 (63.1) |  | 771.2 (64.6) | 771.2 (64.6) |  |
| BMI (Mean ± SD) | 28.5 (7.8) | 28.3 (7.1) | 0.020 | 28.4 (7.7) | 28.4 (7.4) | <0.001 |
| Afib – no. (%) | 682 (39.4) | 1735 (39.3) | 0.002 | 469.0 (39.3) | 469.0 (39.3) | <0.001 |
| CHF – no. (%) | 718 (41.5) | 2331 (52.8) | 0.228 | 529.7 (44.4) | 529.7 (44.4) | <0.001 |
| CKD – no. (%) | 533 (30.8) | 1709 (38.7) | 0.167 | 394.1 (33.0) | 394.1 (33.0) | <0.001 |
| CLD – no. (%) | 139 (8.0) | 269 (6.1) | 0.076 | 89.7 (7.5) | 89.7 (7.5) | <0.001 |
| COPD – no. (%) | 352 (20.3) | 845 (19.1) | 0.030 | 237.5 (19.9) | 237.5 (19.9) | <0.001 |
| CAD – no. (%) | 508 (29.3) | 1968 (44.6) | 0.320 | 396.3 (33.2) | 396.3 (33.2) | <0.001 |
| Stroke – no. (%) | 135 (7.8) | 362 (8.2) | 0.015 | 95.5 (8.0) | 95.5 (8.0) | <0.001 |

Afib=atrial fibrillation; all other abbreviations as in Supplementary Table 1.

Supplementary Table 4. Multivariable analysis for in-hospital mortality using APSIII instead of SOFA before and after propensity score-based overlap weighting.

| Database | Group | Elevated Initial troponin  OR (95% CI) | P value | Elevated Delta troponin  OR (95% CI) | P value |
| --- | --- | --- | --- | --- | --- |
| eICU-CRD | Unweighted | 1.39 (1.24-1.55) | <0.001 | 1.20 (0.94-1.54) | 0.138 |
|  | Overlap weighted | 1.38 (1.22-1.56) | <0.001 | 1.20 (0.94-1.54) | 0.151 |
| MIMIC-III | Unweighted | 1.23 (1.05-1.44) | 0.012 | 1.11 (0.75-1.65) | 0.596 |
|  | Overlap weighted | 1.35 (1.15-1.59) | <0.001 | 1.21 (0.81-1.80) | 0.358 |
| MIMIC-IV | Unweighted | 1.30 (1.12-1.51) | <0.001 | 1.29 (0.97-1.71) | 0.077 |
|  | Overlap weighted | 1.34 (1.15-1.56) | <0.001 | 1.40 (1.04-1.88) | 0.025 |
| Overall | Unweighted | 1.29 (1.19-1.41) | <0.001 | 1.21 (1.02-1.42) | 0.027 |
|  | Overlap weighted | 1.33 (1.22-1.44) | <0.001 | 1.26 (1.06-1.49) | 0.008 |

Supplementary Table 5. Multivariable analysis for 1-year mortality using APSIII instead of SOFA before and after propensity score-based overlap weighting in MIMIC-III.

|  | Unweighted (95% CI) | | overlap weighted (95% CI) | |
| --- | --- | --- | --- | --- |
| Odds ratio | 1.14 (0.96-1.34) | p=0.125 | 1.21 (1.03-1.43) | p=0.023 |
| Hazard ratio | 1.25 (0.97-1.61) | p=0.088 | 1.33 (1.07- 1.67) | p=0.011 |
